# A Combined Deep CNN-LSTM Network for the Detection of Novel Coronavirus (COVID-19) Using X-ray Images

**DOI:** 10.1101/2020.06.18.20134718

**Authors:** Md. Zabirul Islam, Md. Milon Islam, Amanullah Asraf

## Abstract

Nowadays automatic disease detection has become a crucial issue in medical science with the rapid growth of population. Coronavirus (COVID-19) has become one of the most severe and acute diseases in very recent times that has been spread globally. Automatic disease detection framework assists the doctors in the diagnosis of disease and provides exact, consistent, and fast reply as well as reduces the death rate. Therefore, an automated detection system should be implemented as the fastest way of diagnostic option to impede COVID-19 from spreading. This paper aims to introduce a deep learning technique based on the combination of a convolutional neural network (CNN) and long short-term memory (LSTM) to diagnose COVID-19 automatically from X-ray images. In this system, CNN is used for deep feature extraction and LSTM is used for detection using the extracted feature. A collection of 421 X-ray images including 141 images of COVID-19 is used as a dataset in this system. The experimental results show that our proposed system has achieved 97% accuracy, 91% specificity, and 100% sensitivity. The system achieved desired results on a small dataset which can be further improved when more COVID-19 images become available. The proposed system can assist doctors to diagnose and treatment the COVID-19 patients easily.

## 1 Introduction

Coronavirus epidemic that spreads across the whole world has made all the process of each sector lockdown. According to World Health Organization latest estimates, more than seven million people around have been infected and close to 414,060 deaths so far (as of 10^th^ June 2020) [1]. The health system has reached the point of failure even in developed countries, due to the shortage of intensive care units (ICU). COVID-19 patients with worse conditions are filling in ICU. The strain that began to spread in Wuhan of China, is identified from two different coronaviruses, i.e. severe acute respiratory syndrome (SARS) and Middle East respiratory syndrome (MERS) [2]. Symptoms for various types of COVID-19 can range from cold to fever, shortness of breath, and acute respiratory symptoms [3]. In comparison to SARS, the respiratory system is affected by coronavirus as well as kidneys and liver [4].

Coronavirus detection at an early age plays a vital role to control COVID-19 due to the high transmissibility. The diagnosis of coronavirus by gene sequencing for respiratory or blood samples should be confirmed as the main pointer for reverse transcription-polymerase chain reaction (RT-PCR) according to the guidelines of the Chinese government [5]. The process of RT-PCR takes 4-6 hours to get results, which consumes a long time compared to COVID-19’s rapid spread rate. The RT-PCR test kits are in huge shortage, in addition to inefficiency [6]. As a result, many infected cases cannot be detected on time and tend to unconsciously infect others. With the detection of this disease at an early stage, the prevalence of COVID-19 disease will decrease [7]. To alleviate the inefficiency and scarcity of current COVID-19 tests, a lot of effort has been placed into looking for alternative test methods. Another visualization method is to diagnose COVID-19 infections using radiological images such as X-ray images or Computed Tomography (CT). Earlier works show that the anomalies are found in COVID-19 patients in the radiological images in the shape of ground-glass opacities [8]. The researchers claim that a system based on chest radiology can be an important method for identifying, quantifying, and monitoring of COVID-19 events [9]. Today, various researchers from all over the world are working to fight against COVID-19.

Many researchers demonstrate various approaches to detect COVID-19 utilizing X-ray images. Recently, computer vision [10], machine learning [11], [12], [13], and deep learning [14], [15] have been used to automatically diagnose several diverse ailments in the human body that ensures smart healthcare [16], [17]. The deep learning method is used as a feature extractor that enhances the classification accuracies [18]. The detection of tumor forms and area in the lungs, X-ray bone suppression, diagnosis of diabetic retinopathy, prostate segmentation, diagnosis of skin lesions, the examination of the myocardium in coronary CT are the examples of the contributions [19], [20] of deep learning.

Therefore, this paper aims to propose a deep learning based system that combined the CNN-LSTM network to automatically detect COVID-19 from X-ray images. The proposed system, for feature extraction, CNN is used and the LSTM is used to classify COVID-19 based on these features. The 2D CNN and LSTM layout feature combination improve the classification greatly. The dataset used for this paper is collected from multiple sources and a preprocessing performed in order to reduce the noise.

In the following, the contributions of this research are summarized.

a. Developing altogether a combined deep CNN-LSTM network to automatically assist the early diagnosis of patients with COVID-19 efficiently.
b. To detect COVID-19 using chest X-ray, a dataset is formed comprising 421 images.
c. A detailed experimental analysis of the performance of the proposed system was provided in terms of confusion matrix, accuracy, sensitivity, specificity, and f1-score.

The paper is organized as follows: A review of recent scholarly works related to this study is described in Section 2. A description of the overall proposed system with dataset collection and preparation is presented in Section 3. The experimental results and comparative performance of the proposed deep learning system are provided in Section 4. The discussion is demonstrated in Section 5. Section 6 concludes the paper.

## 2 Related Works

With the epidemic of COVID-19, greater attempts have been done to evolve deep learning techniques to diagnose COVID-19 based on clinical images including Computer Tomography (CT) scans and X-rays in the chest. In this literature, a detailed description of recently developed systems applying deep learning techniques in the field of COVID-19 detection has outlined.

Rahimzadeh et al. [21] developed a concatenated CNN for classifying COVID-19 cases based on Xception and ResNet50V2 models using chest X-ray. The developed system used a dataset contained 180 images with COVID-19 patients, 6054 images of pneumonia patients, and 8851 images from normal people. This work selected 633 images in each phase for training and used eight phases. The experimental outcome has obtained 99.56% accuracy and 80.53% recall for COVID-19 cases. Alqudah et al. [22] used artificial intelligence techniques in developing a system to detect COVID-19 from chest X-ray. The used images were classified using deferent machine learning techniques such as support vector machine (SVM), CNN, and Random Forest (RF). The system has obtained 95.2% accuracy, 100% specificity, and 93.3% sensitivity. Loey et al. [23] introduced a Generative Adversarial Network (GAN) with deep learning to diagnose COVID-19 from chest X-ray. The scheme used three pre-trained models named as Alexnet, Googlenet, and Restnet18 for coronavirus detection. The collected data includes 69 images of COVID-19 cases, 79 images of pneumonia bacterial cases, 79 images of pneumonia virus cases, and 79 images of normal cases. The Googlenet was selected as a main deep learning technique with 80.6% test accuracy in four classes scenario, Alexnet with 85.2% test accuracy in three classes scenario, and Googlenet with 99.9% test accuracy in two classes scenario.

Ucar et al. [24] proposed a COVID-19 detection system based on deep architecture from X-ray images. In the developed system, the dataset includes 76 images of COVID-19 cases, 4290 images of pneumonia cases, and 1583 images of normal cases. The scheme has achieved 98.3% accuracy for COVID-19 cases. Apostolopoulos et al. [25] introduced a transfer learning strategy with CNN for the diagnosis of COVID-19 cases automatically by extracting the essential features from the chest X-ray. The system used five variants of CNN named VGG19, Inception, MobileNet, Xception, and Inception-ResNetV2 to classify COVID-19 images. In the developed system, the dataset includes 224 images of COVID-19 patients, 700 images of pneumonia patients, and 504 images of normal patients. The dataset is split using the concept of ten-fold cross-validation for training and evaluation purposes. The VGG19 was selected as a main deep learning model with 93.48% accuracy, 92.85% specificity, and 98.75% sensitivity in the developed system. Bandyopadhyay et al. [26] proposed a novel model that combined the LSTM-GRU to classify the confirmed, released, negative, and death cases on COVID-19 automatically. The developed scheme achieved 87% accuracy on the confirmed case, 67.8% accuracy on the negative case, 62% accuracy on the deceased case, and 40.5% accuracy on the released case for the prediction of COVID-19. Khan et al. [27] presented a deep learning network to predict COVID-19 cases automatically from chest X-ray. In the developed system, the dataset includes 284 images of COVID-19 cases, 330 images of pneumonia bacterial cases, 327 images of pneumonia viral cases, and 310 images of normal cases. The proposed system has obtained an overall 89.5% accuracy, 97% precision, and 100% recall respectively for COVID-19 cases.

Kumar et al. [28] introduced a deep learning methodology for the classification of COVID-19 infected patients using chest X-ray. The scheme used nine pre-trained models for the features extraction and support vector machine for the classification. They prepared two sets of datasets containing 158 X-ray images for both COVID-19 and non-COVID-19 patients. The ResNet50 plus SVM is statistically superior to other models with accuracy and the f1-score was 95.38%, 95.52% respectively. Horry et al. [29] developed a system based on pre-trained model to detect COVID-19 from chest X-ray. They used Xception, VGG, Resnet, and Inception for the classification of COVID-19 patients. The dataset used in the system included 115 images of COVID-19 cases, 322 images of pneumonia cases, and 60361 images of normal cases. The results of around 80% both for recall and of precision both for VGG16 and VGG19 classifiers are appraised. Hemdan et al. [30] introduced a deep learning technique to detect COVID-19 using X-ray images. The framework includes seven pre-trained models such as VGG19, MobileNetV2, InceptionV3, ResNetV2, DenseNet201, Xception, and InceptionRes-NetV2. In the developed system, 50 X-ray images are considered where 25 images from COVID-19 infected patients and the rest 25 images from non-COVID-19 patients. Among all tested classifiers, VGG19 and DenseNet201 models achieved the highest values of 90% accuracy with 83% precision.

## 3 Methods and Materials

The overall system for the detection for COVID-19 requires several phases, which are shown in Fig. 1. At first, raw X-ray images are passing through the preprocessing pipeline. Data resizing, shuffling, normalization, and augmentation process was done in the preprocessing pipeline. Then, the preprocessed dataset is partitioned into a training set and testing set. Afterward, we have trained CNN, and CNN-LSTM architecture using the training dataset of the workflow. After each epoch, the training accuracy and loss are found in the proposed system. At the same time, using 5-fold cross-validation the validation accuracy and loss are also obtained. The performance is measured of some evaluation metrics such as confusion matrix, accuracy, specificity, sensitivity, and f1-score of the proposed system.

**Fig. 1.**
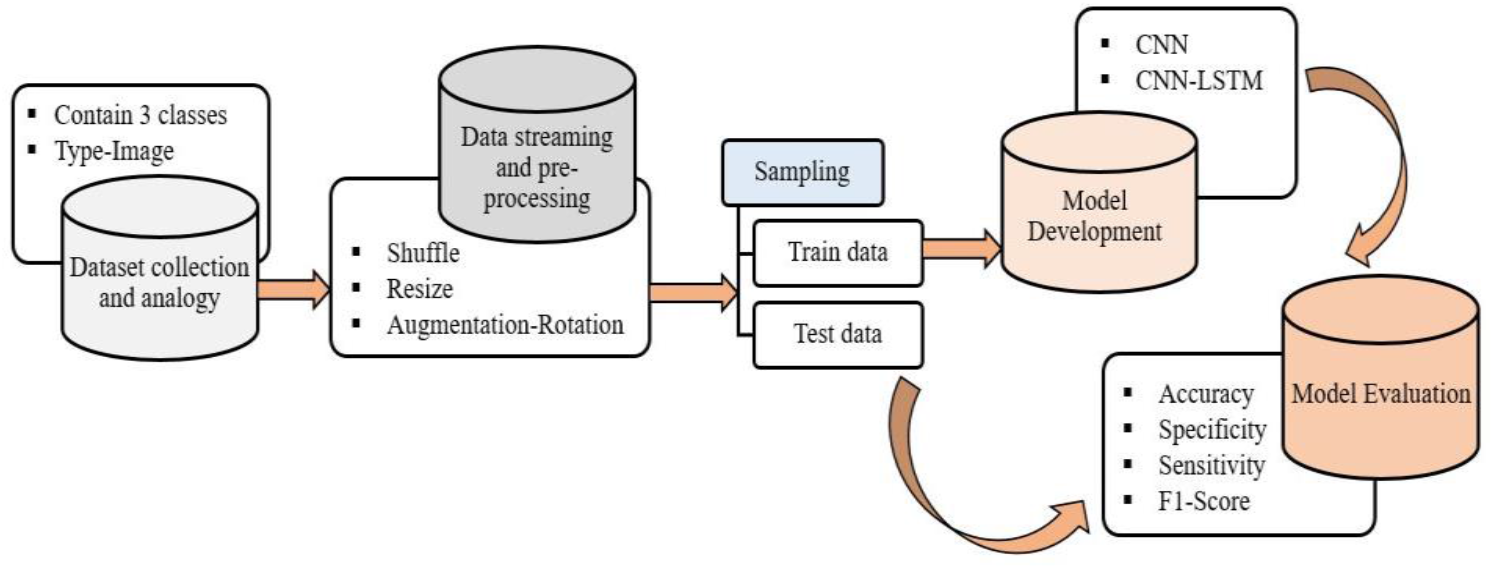
The overall system architecture of the proposed COVID-19 detection system

### 3.1 Dataset Collection and description

As the emergence of COVID-19 being very recent, none of these large repositories contains any COVID-19 labeled data, thereby requiring us to rely on at least two datasets for normal, pneumonia, and COVID-19 source images. Firstly, 141 X-ray images of COVID-19 cases are collected from GitHub developed by Joseph Paul Cohen [31]. Secondly, 140 X-ray images of pneumonia cases and 140 X-ray images of normal cases are collected from the Kaggle repository [32]. The main objective of this dataset selection is to make it available to the public so that it is accessible and extensible to researchers. Further studies may be more efficient to diagnose COVID-19 patients based on this dataset. We resized the images to 224 × 224 pixels. The number of X-ray images of each set is partitioned in Table 1. The visualization of X-ray images of each class is shown in Fig. 2.

**Table 1.** The partitioning description of used dataset

| Data/Cases | COVID-19 | Normal | Pneumonia | Overall |
| --- | --- | --- | --- | --- |
| Training | 119 | 119 | 119 | 357 |
| Testing | 22 | 21 | 21 | 64 |
| Overall | 141 | 140 | 140 | 421 |

**Fig. 2.**
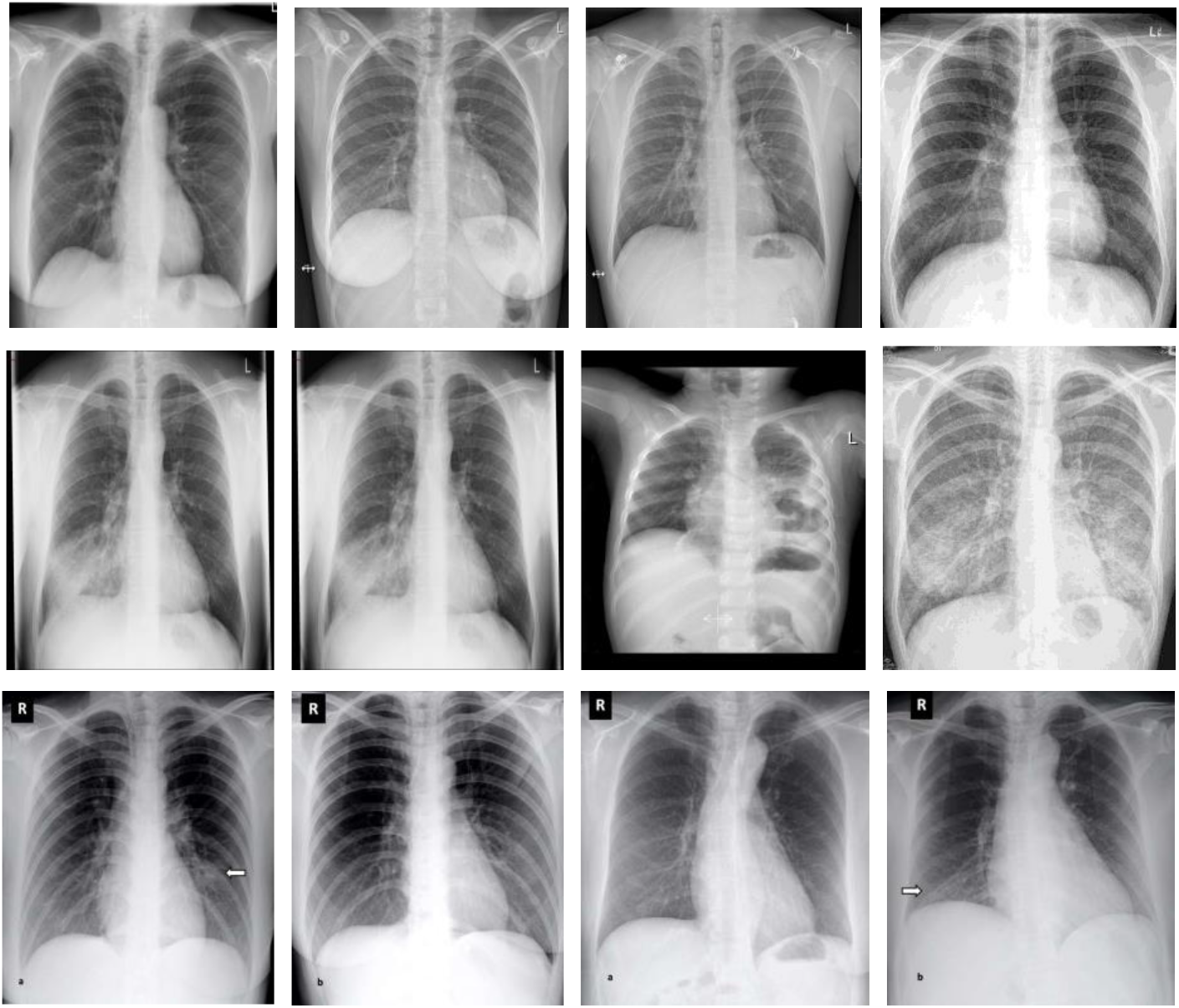
The images in the first row show 4 sample images COVID-19 cases. The images in the second row are 4 sample images of pneumonia cases and the third row are 4 sample images of normal cases.

### 3.2 Development of Combined Network

The proposed architecture is formed with the combination of CNN and LSTM network which is briefly described as follows.

#### 3.2.1 Convolutional Neural Network

A particular type of multilayer perceptron is a convolutional neural network. However, a simple neural network can not learn complex features but a deep architecture network can learn it. On many applications [33], [34] such as image classification, object detection, and medical image analysis, CNN’s have shown excellent performance. The main idea behind CNN is that it can obtain local features from higher layer inputs and transfer them into lower layers for more complex features. CNN is comprised of a convolutional layer, pooling layer, and fully connected (FC) layer. A typical CNN architecture along with these layers is depicted in Fig. 3.

**Fig. 3.**
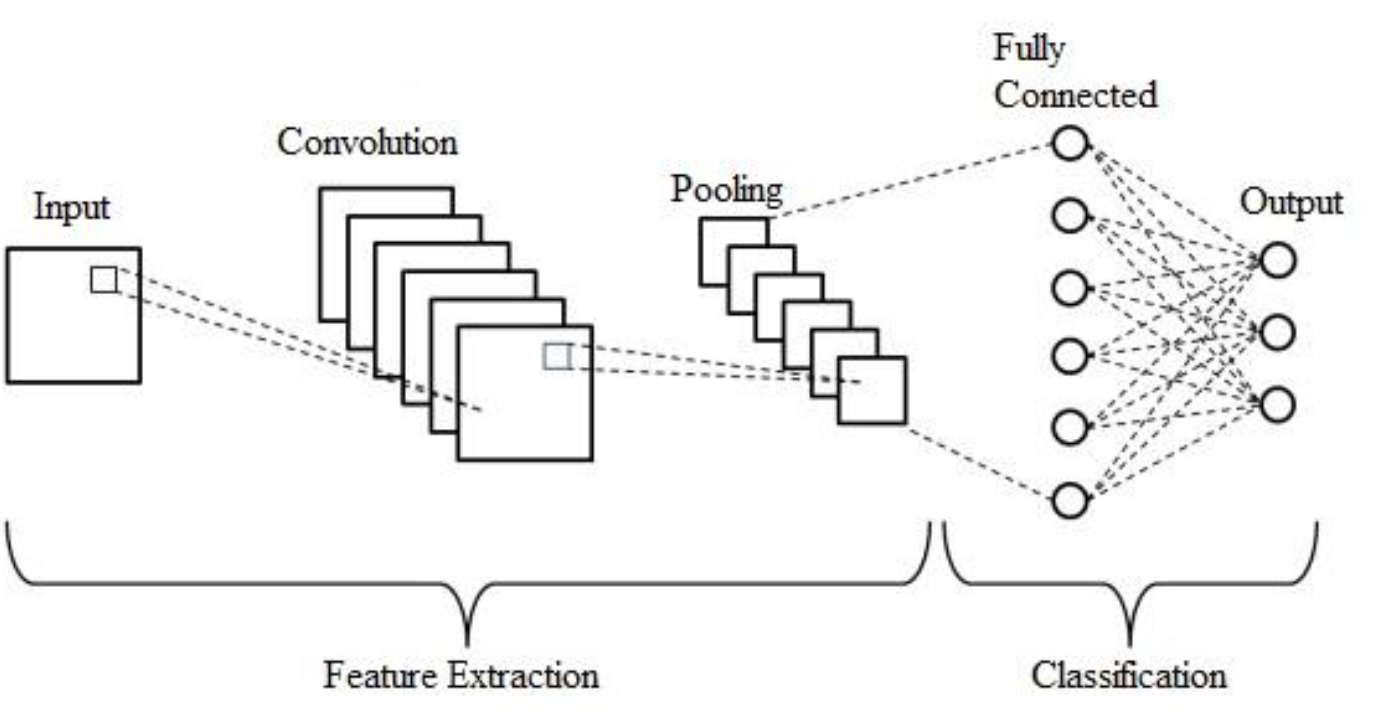
A typical architecture of the convolutional neural network.

The convolutional layer includes a set of kernels [35] for determining a tensor of feature maps. These kernels convolve entire input using “stride(s)” so that the dimensions of the output volume become integers [36]. The input volume’s dimensions reduce after the convolutional layer for the striding process. Therefore, zero-padding [37] is required to pad the input volume with zero’s to maintain the dimension of the input volume with low-level features. The operation of convolution layer is given as:

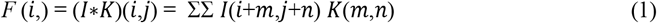

where I refers to the input matrix, K refers to a 2D filter of size m × n, and F refers output of the 2D feature map. The operation of the convolution layer is denoted by I*K.

For increasing the nonlinearity in the feature maps, the rectified linear unit (ReLU) layer is used [38]. ReLU computes the activation by keeping the threshold the input at zero. It is mathematical expression is given as:

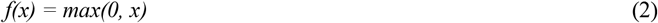

The pooling layer [39] is performed downsampling of the given input dimension to reduce the number of parameters. Max Pooling is the most common method which produces the maximum value in the input region. The fully-connected layer [40] is used as a classifier to make a decision based on obtained features from convolution and pooling layers. A full CNN architecture is developed with these layers which are discussed above.

#### 3.2.2 Long Short-Term Memory

Long short-term memory is an improvement from the recurrent neural network (RNN). LSTM proposes memory blocks instead of conventional RNN units to solve vanishing and exploding gradient problem [41]. LSTM added a cell state to save long-term states which is the main difference from RNN. LSTM network can remember and connect the previous information to the present [42]. The structure of the LSTM network is shown in Fig. 4. The LSTM is combined with three gates such as the input gate, the forget gate and the output gate where *x_t_* refers the current input, *C*_t_ and *C_t_−*1 refer the new and previous cell state respectively, *h_t_* and *h_t_−*1 refer the current and previous output respectively.

**Fig. 4.**
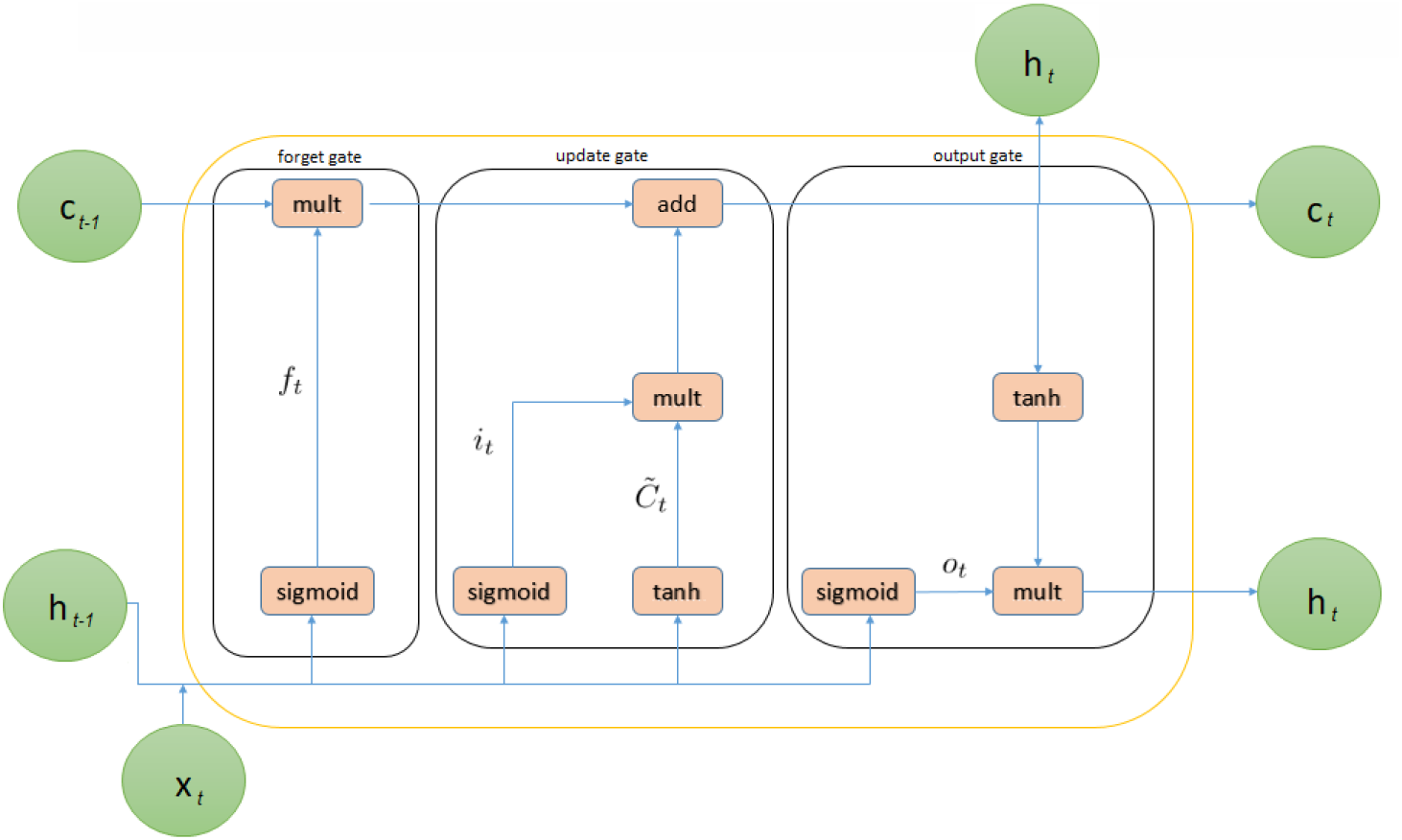
The internal structure of Long short-term memory.

The principle of the input gate of the LSTM is shown in the following forms.

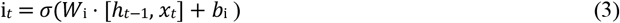

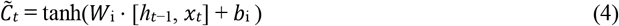

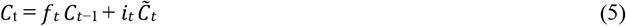

where (3) is used to pass *h_t_−*1 and *x_t_* through the sigmoid layer to decide which portion of information should be added. Subsequently, (4) obtains new information after passing *h_t_−*1 and *x_t_* through the tanh layer. The current moment information 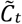 and long-term memory information *C_t_−*1 into *C*_t_ are combined in (5), where *it* refers to sigmoid output, and 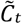 refers to tanh output. Here, Wi refers to weight matrices and bi refers to input gate bias of LSTM. Then, the LSTM’s forget gate allows the selective passage of information using a sigmoid layer and a dot product. The decision about whether to forget from the previous cell’s related information with a certain probability is done using (6) where Wf refers to the weight matrix, b_f_ is the offset, and σ refers to the sigmoid function.

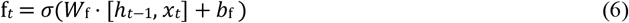

The LSTM’s output gate determines the states which are required to be continued by the *h_t_−*1 and *x_t_* inputs following to (7) and (8). The final output is obtained to multiply with the state decision vectors passing new information Ct through the tanh layer.

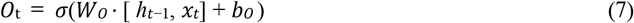

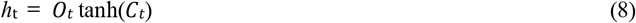

where *W*o and *b*o are the output gate’s weighted matrices and the output gate’s bias of LSTM respectively.

#### 3.2.3 Combined CNN-LSTM Network

In this paper, a combined method was developed to automatically detect the COVID-19 cases using three types of X-ray images. The structure of this architecture is developed by combining CNN and LSTM networks where CNN is used to extract the complex features from images and LSTM is used as a classifier.

Fig. 5 illustrates the proposed hybrid network for COVID-19 detection. Each convolution block combined with two or three 2D CNN and one pooling layer; the rectified linear units are activated as an activation function. The convolution kernel is extracted convolution feature by multiplying the superposition matrix in all convolution operations. The maximum-pooled filter of the feature map is used for feature extraction after a two-dimensional convolution, and the filter’s step size is two. In the last part of the architecture, the function map is transferred to the LSTM layer to extract time information. Finally, the fully connected layer of the softmax function is used to predict into three categories (COVID-19, pneumonia, and normal).

**Fig. 5.**
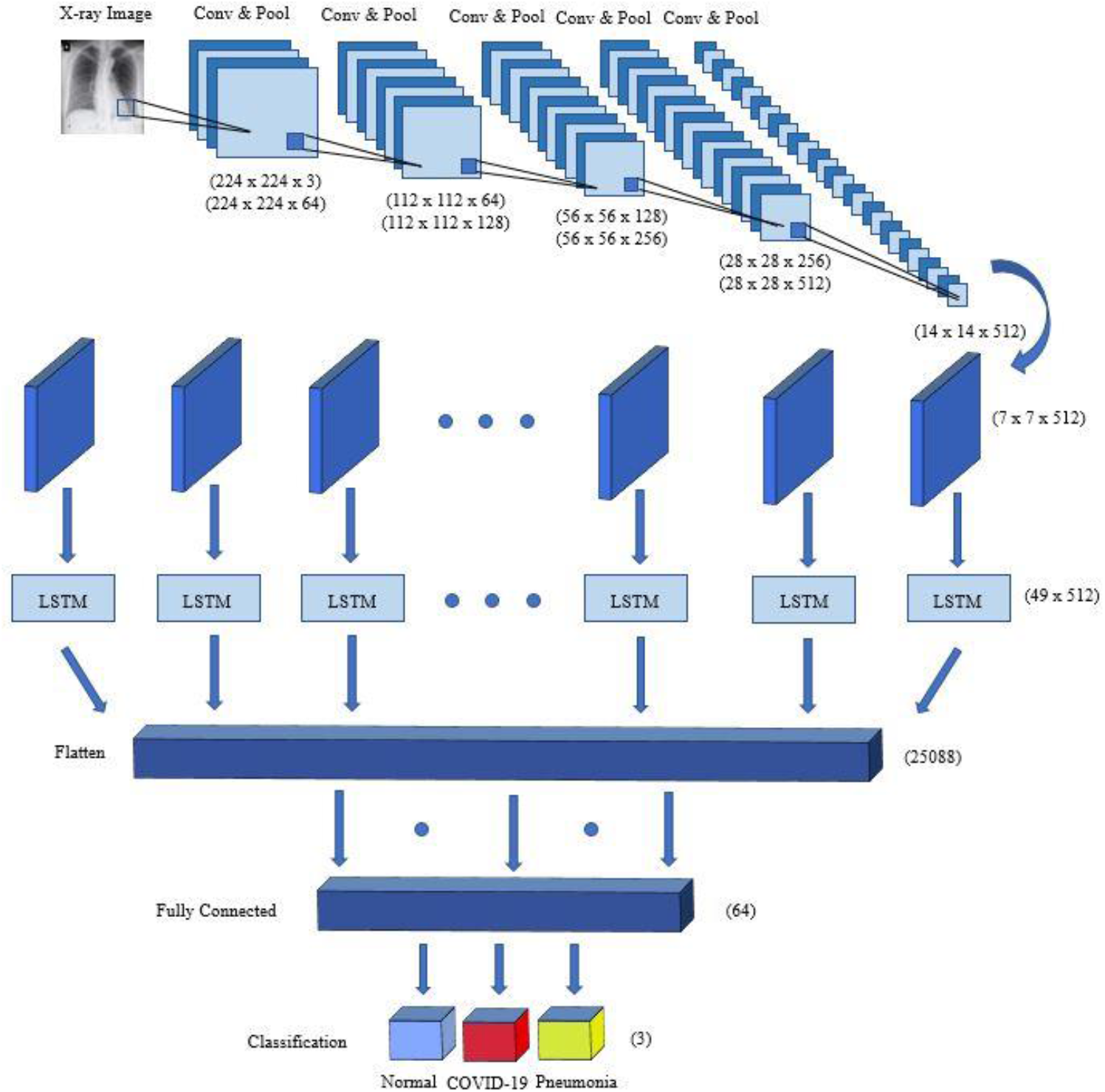
An illustration of the proposed hybrid network for COVID-19 detection

The structure of the proposed architecture is shown in Table 2. From Table 2, it is found that the Layers 1–17 of the network are convolutional layers, and layer 18 is the LSTM layer. Finally, a fully connected layer is added for predicting the output. After the pooling layer, the output shape is found (none, 7, 7, 512). The input size of the LSTM layer is (49, 512). After analyzing LSTM time characteristics, finally, the architecture sorts X-ray images through a fully connected layer.

**Table 2.** The full summary of CNN-LSTM network

| Layer | Type | Kernel Size | Stride | Kernel | Input Size |
| --- | --- | --- | --- | --- | --- |
| 1 | Convolution2D | $3 \times 3$ | 1 | 64 | $224 \times 224 \times 3$ |
| 2 | Convolution2D | $3 \times 3$ | 1 | 64 | $224 \times 224 \times 64$ |
| 3 | Pool | $2 \times 2$ | 2 | - | $224 \times 224 \times 64$ |
| 4 | Convolution2D | $3 \times 3$ | 1 | 128 | $112 \times 112 \times 64$ |
| 5 | Convolution2D | $3 \times 3$ | 1 | 128 | $112 \times 112 \times 128$ |
| 6 | Pooling | $2 \times 2$ | 2 | - | $112 \times 112 \times 128$ |
| 7 | Convolution2D | $3 \times 3$ | 1 | 256 | $56 \times 56 \times 128$ |
| 8 | Convolution2D | $3 \times 3$ | 1 | 256 | $56 \times 56 \times 256$ |
| 9 | Pool | $2 \times 2$ | 2 | - | $56 \times 56 \times 256$ |
| 10 | Convolution2D | $3 \times 3$ | 1 | 512 | $28 \times 28 \times 256$ |
| 11 | Convolution2D | $3 \times 3$ | 1 | 512 | $28 \times 28 \times 512$ |
| 12 | Convolution2D | $3 \times 3$ | 1 | 512 | $28 \times 28 \times 512$ |
| 13 | Pool | $2 \times 2$ | 2 | - | $28 \times 28 \times 512$ |
| 14 | Convolution2D | $3 \times 3$ | 1 | 512 | $14 \times 14 \times 512$ |
| 15 | Convolution2D | $3 \times 3$ | 1 | 512 | $14 \times 14 \times 512$ |
| 16 | Convolution2D | $3 \times 3$ | 1 | 512 | $14 \times 14 \times 512$ |
| 17 | Pool | $2 \times 2$ | 2 | - | $14 \times 14 \times 512$ |
| 18 | LSTM | - | - | - | $49 \times 512$ |
| 19 | FC | - | - | 64 | 25,088 |
| 20 | Output | - | - | 3 | 64 |

### 3.3 Performance Evaluation Metrics

The following metrics are used to measure the performance of the proposed system. It is noted that TP is the correctly predicted COVID-19 cases, FP is the normal or pneumonia cases that were misclassified as COVID-19 by the proposed system, TN is the normal or pneumonia cases that were correctly classified, while the FN refers to COVID-19 cases that were misclassified as normal or pneumonia cases.

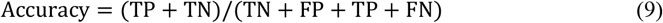

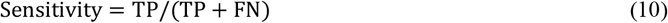

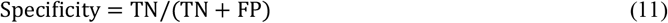

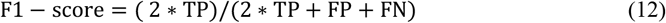

## 4 Experimental Results Analysis

### 4.1 Experimental Setup

In the experiment, the dataset was split into 85% and 15% for training and testing respectively. Using k-fold cross-validation, results were obtained according to 5 different k values such as (k=1-5). The proposed network consists of twelve layers as described in Table 2, the learning rate is 0.0001, and the maximum epochs number is 200 which were determined experimentally. The CNN and CNN-LSTM networks have been implemented using Python and the Keras package with TensorFlow2 on Intel(R) Core(TM) i7-2.2 GHz processor. Besides, the experiments were executed using the graphical processing unit (GPU) NVIDIA GTX 1050 Ti and RAM with 4 GB and 16GB, respectively.

### 4.2 Results Analysis

Fig. 6 depicts the confusion matrix of the test phase of the proposed CNN-LSTM and the competitive CNN architecture for COVID-19 disease classification. Though both architectures classify all 22 images of COVID-19 perfectly, there are four misclassified images for CNN and two misclassified images for CNN-LSTM among 64 images. It is found that the proposed CNN-LSTM network outperforms the competitive CNN network as it has better, and consistent true positive and true negative values and lesser false-negative and false-positive values. Hence, the proposed system can efficiently classify COVID-19 cases.

**Fig. 6.**
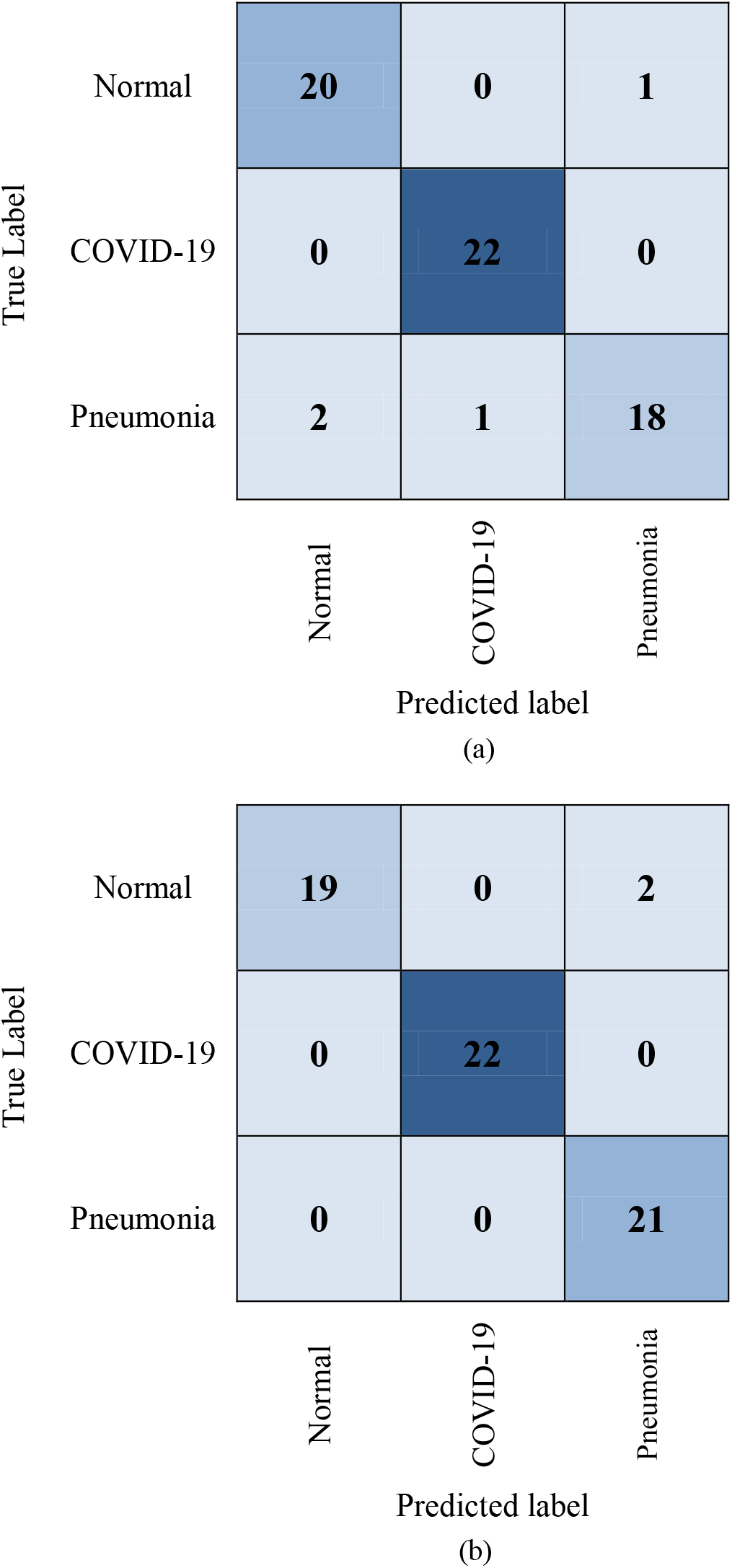
Confusion matrix of the proposed COVID-19 detection system. (a) CNN (b) CNN-LSTM

Moreover, Fig. 7 illustrates the performance evaluation of the CNN classifier graphically with accuracy and cross-entropy (loss) in the training and validation phase. The training and validation accuracy is found at 92% and 95% respectively. Similarly, the training and validation loss are found 0.18 and 0.09 respectively for the CNN architecture.

**Fig. 7.**
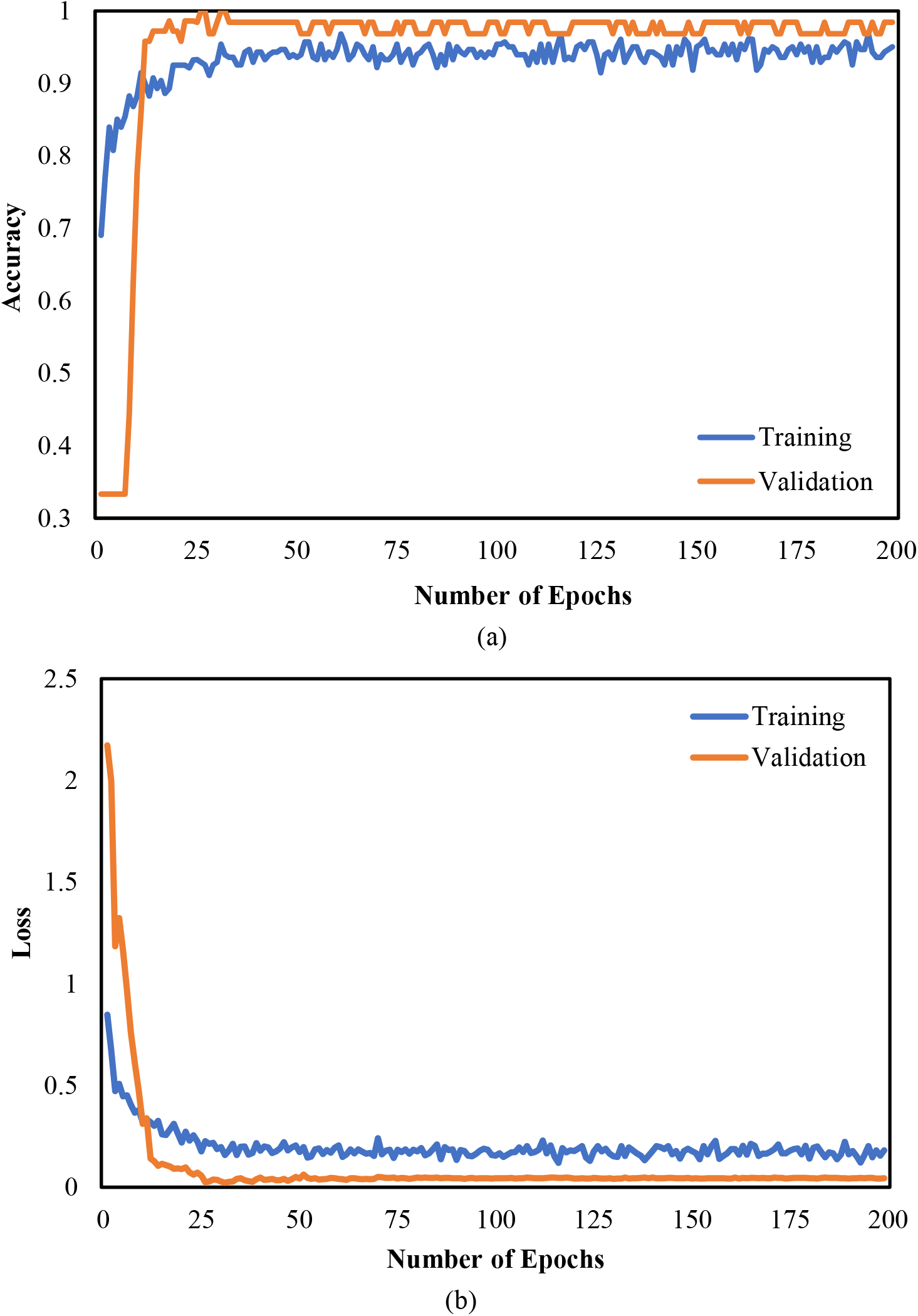
Evaluation metrics of COVID-19 detection system based on CNN architecture (a) Accuracy (b) Loss

Further, Fig. 8 depicts the performance evaluation of the CNN-LSTM classifier graphically with accuracy and cross-entropy (loss) in the training and validation phase. The training and validation accuracy is found 95% and 98% respectively. Similarly, the training and validation loss are found 0.18 and 0.04 respectively for the CNN-LSTM architecture. The best scores of training and validation accuracy were achieved for the CNN-LSTM architecture comparing with the CNN architecture.

**Fig. 8.**
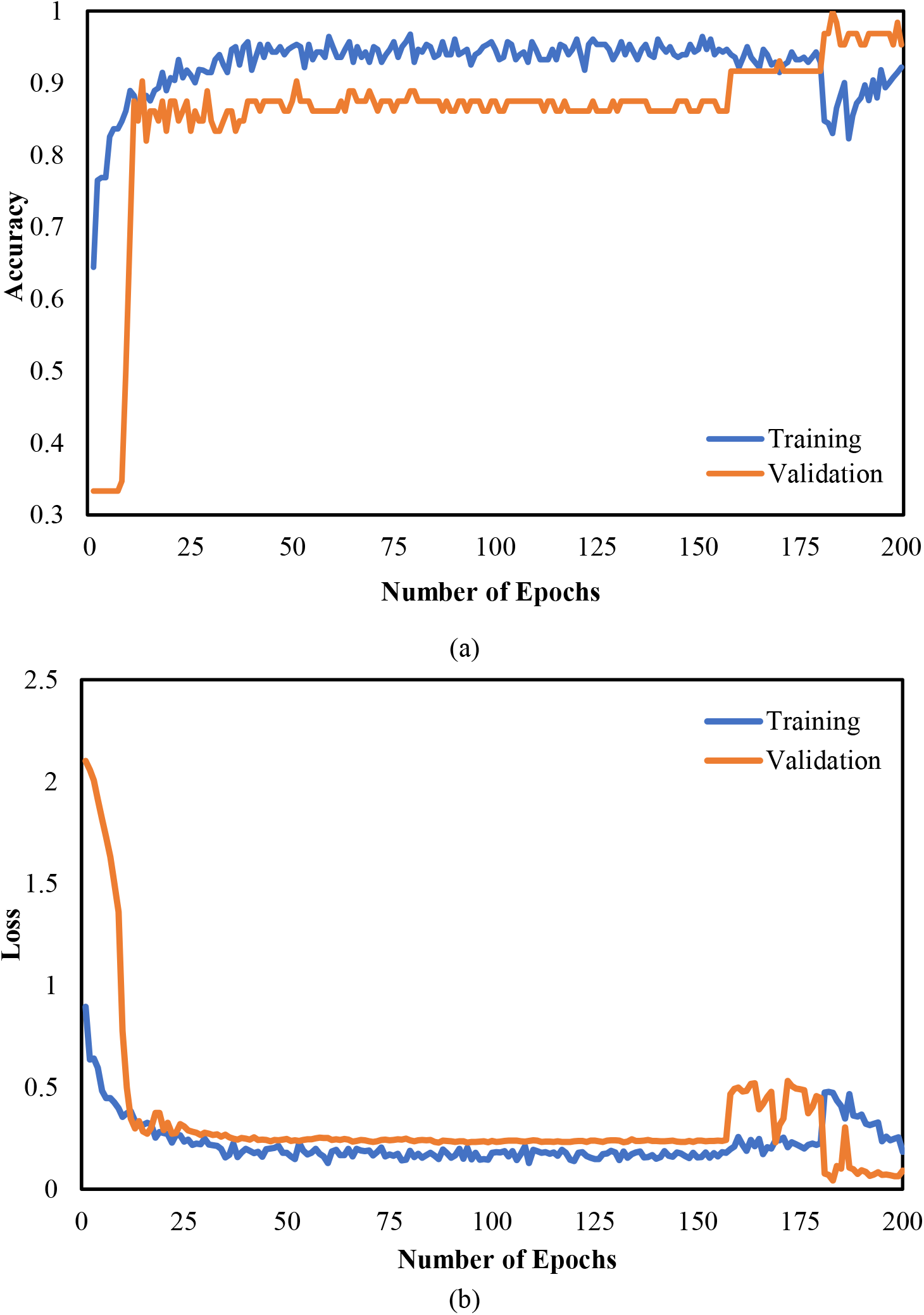
Evaluation metrics of COVID-19 detection system based on CNN-LSTM architecture (a) Accuracy (b) Loss

The overall accuracy, specificity, sensitivity, and f1-score for each case of CNN architecture are summarized in Table 3 and visually shown in Fig. 9. The obtained accuracy is 94% for COVID-19, pneumonia, and normal cases. The specificity is found 96%, 95%, and 91% for COVID-19, pneumonia, and normal cases respectively. The sensitivity has achieved 100%, 86%, and 95% for COVID-19, pneumonia, and normal cases respectively. The f1-score is obtained 98%, 90%, 93% for COVID-19, pneumonia, and normal cases respectively. While the highest specificity, sensitivity, and f1-score are obtained by COVID-19, the lower values of sensitivity and f1-score are found in pneumonia cases.

**Table 3.** Performance of the CNN network

| Class | Accuracy (%) | Specificity (%) | Sensitivity (%) | F1-Score (%) |
| --- | --- | --- | --- | --- |
| COVID-19 | 0.94 | 0.96 | 1.00 | 0.98 |
| Pneumonia | 0.94 | 0.95 | 0.86 | 0.90 |
| Normal | 0.94 | 0.91 | 0.95 | 0.93 |

**Fig. 9.**
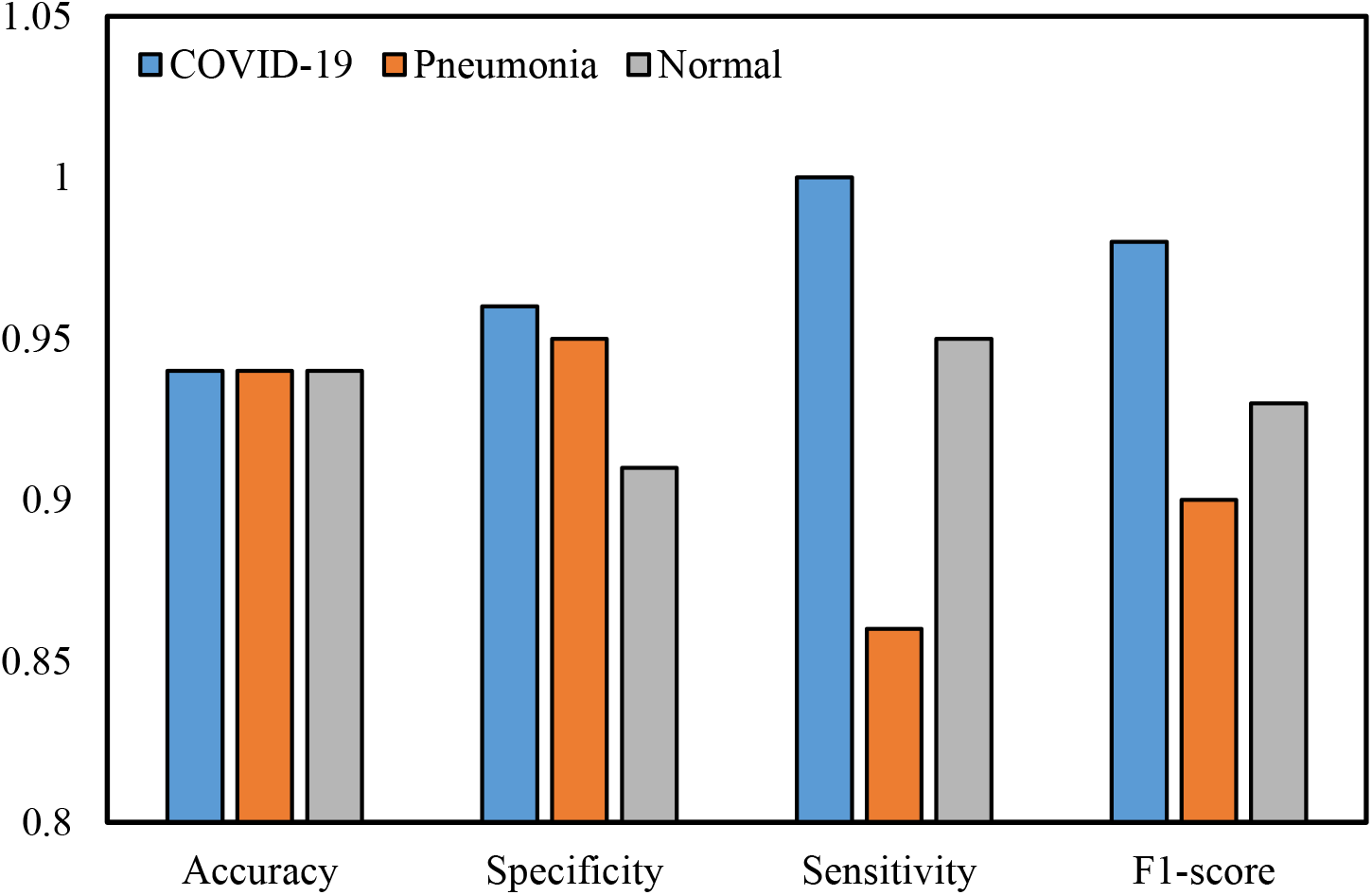
The graphical representation of the results of the CNN network

Furthermore, Table 4 and Fig. 10 shows the performance metrics of each class of the developed CNN-LSTM network. The accuracy is 97% for COVID-19, pneumonia, and normal cases. The specificity is obtained 91%, 100%, and 100% for COVID-19, pneumonia, and normal cases respectively. The sensitivity is achieved 100% for both COVID-19 and pneumonia cases and 90% for normal cases. The f1-score is found 100% for COVID-19 and 95% for both pneumonia, and normal cases. While the maximum sensitivity and f1-score are achieved by COVID-19, the lower values of sensitivity and f1-score are obtained in normal and pneumonia cases respectively.

**Table 4.** Performance of the CNN-LSTM network

| Class | Accuracy (%) | Specificity (%) | Sensitivity (%) | F1-score (%) |
| --- | --- | --- | --- | --- |
| COVID-19 | 0.97 | 0.91 | 1.00 | 1.00 |
| Pneumonia | 0.97 | 1.00 | 1.00 | 0.95 |
| Normal | 0.97 | 1.00 | 0.90 | 0.95 |

**Fig. 10.**
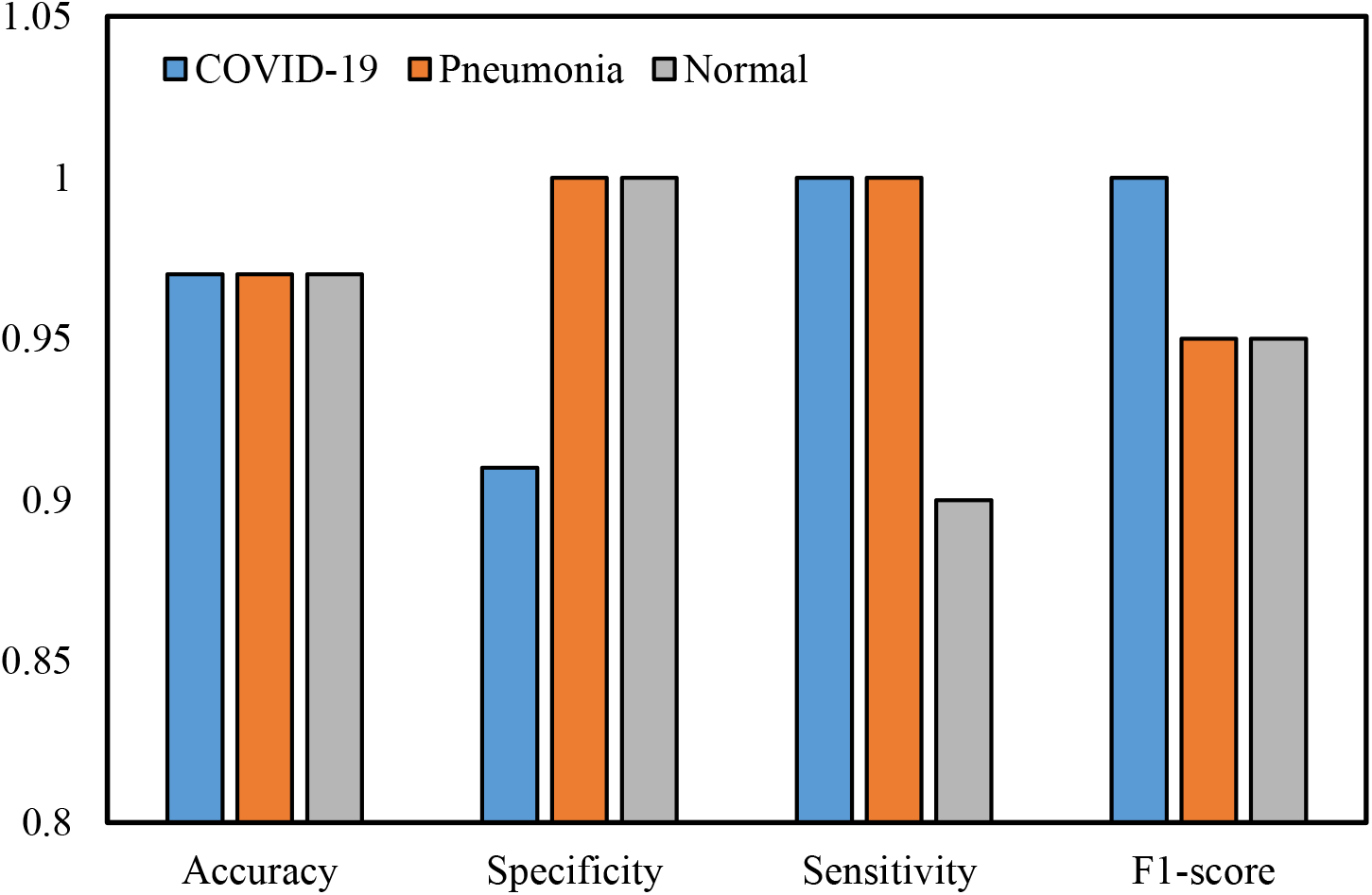
The graphical representation of the results of the CNN-LSTM network

From the experimental findings, it is evident that the CNN architecture achieved 94% accuracy, 96% specificity, and 100% sensitivity after experimental verification for the COVID-19 infected cases. Comparing the outcomes, the proposed CNN-LSTM network achieved an overall 97% accuracy, 91% specificity, and 100% sensitivity respectively for the COVID-19 cases. The main purpose of this research is to achieve good results in detecting COVID-19 cases and not detecting false COVID-19 cases. Hence, experimental results reveal that the proposed CNN-LSTM architecture outperforms competitive CNN network.

## 5. Discussions

By analyzing the results, it is demonstrated that a combination of CNN and LSTM has significant effects on the detection of COVID-19 based on the automatic extraction of features from X-ray images. The proposed system could distinguish COVID-19 from pneumonia and normal cases with high accuracy. A comparative study of systems with our proposed CNN-LSTM system is shown in Table 5. From Table 5, it is found that some of the proposed systems [23], [27], [30], and [21] obtained a slightly lower accuracy of 80.6%, 89.5%, 90%, and 91.4% respectively. The moderately highest accuracy of 93.5%, 95.2%, and 95.4% are found at [25], [22], and [28] respectively. The result of our proposed system is superior compared to other existing systems. The existing systems achieved accuracy between 80.6%-95.4% which slightly less than our proposed system.

**Table 5.** Comparison of the proposed CNN-LSTM architecture with existing systems in terms of accuracy

| Author | Architecture | Accuracy (%) |
| --- | --- | --- |
| Rahimzadeh et al. [21] | Xception + ResNet50V2 | 91.4 |
| Alqudah et al. [22] | AOCT-NET | 95.2 |
| Loey et al. [23] | Googlenet | 80.6 |
| Apostolopoulos et al. [25] | VGG19 | 93.5 |
| Khan et al. [27] | CoroNet (Xception) | 89.5 |
| Kumar et al. [28] | Resnet50 + SVM | 95.4 |
| Hemdan et al. [30] | VGG19 | 90.0 |
| **Proposed System** | **CNN-LSTM** | **97.0** |

## 6. Conclusion

The COVID-19 cases are increasing daily, many countries face resource shortages. Hence, it is necessary to identify every single positive case during this health emergency. We introduced a deep CNN-LSTM network for the detection of novel COVID-19 from X-ray images. Here, CNN is used as a feature extractor and the LSTM network as a classifier for the detection of coronavirus. The developed system obtained an overall 97% accuracy for all cases and 100% accuracy for COVID-19 cases. The proposed CNN-LSTM and competitive CNN architecture are applied both on the same dataset. The extensive experimental results reveal that the proposed architecture outperforms competitive CNN network. In these global COVID-19 pandemics, we hope that the proposed system would able to develop a tool for COVID-19 patients and reduce the workload of the medical diagnosis for COVID-19.

The limited number of COVID-19 X-ray images is the main limitation of this study which would be overcome if more images are found in the coming days.

## Data Availability

Websites

